# Variation in admissions from hospital emergency departments in the English NHS

**DOI:** 10.1101/2025.10.08.25337601

**Authors:** Martin Chalkley, Nikita Jacob, Rita Santos, Luigi Siciliani

## Abstract

**Background:** Emergency admissions to hospitals via the Emergency Department (ED) are a growing concern internationally due to their impact on healthcare costs. The aim of this study is to document the variation between hospitals in the English National Health Service in respect of their ED admissions before and after taking account of patient casemix and the primary care environment.

**Methods:** Fixed-effect regression analysis of 13,888,084 ED attendances from 9,063,518 patients from all NHS hospitals for 2018/19. The primary outcome was the hospital-specific likelihood of a patient being admitted after an ED attendance. Extensive controls were included for characteristics of the patient, their attendance and their GP practice.

**Results:** On average, 20% of ED attendances resulted in emergency admissions. Significant variation in admission rates was observed across English hospitals, both before and after adjusting for patient and system characteristics. Increased admission likelihood was found for:

- older adults, non-White ethnic groups, and patients from more deprived areas.
- patients arriving by ambulance or during periods when GP practices were closed.
- patients living closer to their GP.
- GP practices for which patients are less aware of extended hours and have higher chronic disease prevalence.

After allowing for controls, considerable unexplained variation in hospital admission rates persisted, ranging from 12% to 35%, with an uneven distribution across the country.

**Conclusion:** There is substantial and persistent variability in emergency admission rates across English NHS hospitals, after controlling for a comprehensive range of patient and system characteristics. This highlights that a uniform approach to managing admissions may be ineffective and that tailored strategies, considering local resources, patient needs, and hospital-specific capabilities, are essential to reduce unnecessary admissions while ensuring equitable access to essential care. Future research should further explore the roles of alternative emergency care services, patient socioeconomic factors, and broader emergency care infrastructure.

**Key messages:** *The state of knowledge:*

- Emergency admissions via the ED are a growing concern internationally due to their impact on costs, hospital crowding, and the potential for subsequent admissions.
- Considerable variation exists in hospital admission rates, even after accounting for patient case mix and the overall healthcare environment.
- Existing studies largely focus on predicting whether a particular patient will be admitted from an ED.
- There are few studies that consider variation across hospitals in ED admissions, and those that do were primarily set in the US healthcare system.

*What this study adds:*

- The first in the UK to examine the variation between hospitals in their ED admission rates, accounting for both patient characteristics, the local healthcare delivery environment, and the specific provision of out-of-hospital services at the patient level.
- It highlights that significant, unexplained variation in admission rates persist among hospitals even after extensive controls for patient demographics, socioeconomic status, attendance characteristics (e.g., arrival by ambulance, time of arrival), and GP practice characteristics.
- It identifies specific patient and GP practice characteristics that are associated with higher admission rates (e.g., older adults, non-white ethnic groups, more deprived areas, arrival by ambulance, arrival when GP practice is closed).
- It provides geographical insights into where higher adjusted admission rates persist (e.g., Liverpool, Bradford, Buckinghamshire, and Guildford).

*How this study might guide future practise and policy:*

- The findings suggest that a uniform national approach to managing emergency admissions is likely to be ineffective.
- Policy and practice should adopt tailored strategies that consider local resources, specific patient needs, and the capabilities and policies of individual hospitals to effectively reduce unnecessary admissions.
- It implies that well-placed urgent care facilities and improved transport options could alleviate pressure on emergency services in certain areas.
- The study informs decision-makers with oversight of healthcare systems on where efforts to reduce admissions could be better targeted and provides a basis for identifying examples of successful admission reduction strategies.
- It points to the need for future research to further explore the roles of alternative emergency care services, patient socioeconomic factors, and the broader emergency care infrastructure.

## Introduction

The growth in admissions to hospitals that follow an emergency department (ED) attendance is a widespread international concern due to the impact of admissions on costs^1^, hospital crowding, and subsequent potential further admissions^2^. Various national interventions are designed to moderate emergency admissions, including changes in hospital payments for admitted patients in the English National Health Service (NHS)^3^. ED admission decisions are clinically driven, but they are also made in the context of hospital capacity, patient complexity, and system-level incentives.

Our study is motivated by the fact that there is considerable variation in the propensity to admit patients across different hospitals. Variation persists even after observable differences in patient case mix and the overall healthcare environment are accounted for. Identifying this residual variation is important because it provides the evidence needed to better target efforts to reduce admissions and can provide examples of where reduction in admissions have already taken place, serving as examples of what works.

A number of existing studies^4–8^ including a survey^9^ have focused on how to predict whether a particular patient will be admitted from an ED. In contrast to those studies our focus is on how hospitals’ admission rates vary, once confounders such as the patient characteristics, their primary care provider and the characteristics of the area that they live in are accounted for. Hence we are interested in what cannot be explained and how that varies across hospitals. This also entails different potential users of our research findings. Whereas studies of the predictors of admission are directed at those managing hospitals, our findings address the concerns of decision-makers with oversight of healthcare systems. We drew on these studies in selecting relevant control variables to isolate hospital fixed effects, in particular the importance of accounting for the day of the week and time of arrival as potential predictors of admission ^6^.

Variation in healthcare delivery across different local areas has been extensively documented and studied for approaching 100 years ^10–12^ but there are only few studies that consider variation across hospitals in ED admissions. Two studies^13,14^ we identified were set in the US healthcare system. The first of these^13^ is concerned with describing variation between hospitals after risk-adjusting their patient mix. In contrast to that approach we risk-adjust not only the patient mix, but account for differences in the out-of-hospital emergency care both at the patient level and the local healthcare system. Closer to our approach the second study^14^ considers hospital-level admission rates and as with our study uses patient level risk-adjusters. Unlike our study however, there are no patient level out-of-hospital measures of health care provision. Hence, this is the first study to examine the variation between hospitals in their ED admission rates accounting for both patient characteristics, the local healthcare delivery environment and the particular provision of out-of-hospital services at the the patient level. Our setting also differs from previous studies of variation because its setting being the NHS in England which is a wholly publicly-funded healthcare system. This is an important distinction from the perspective of policy since there are system-wide interventions that could mitigate high ED admission rates.

### Research Methods

#### Data Sources

We use anonymised attendance-level data from all consultant-led 24-hour ED National Health Service (NHS) hospitals in England in 2018/19. These data are routinely collected and reported in a number of datasets for the English NHS, specifically the Accident and Emergency (A&E) and Admitted Patient Care (APC) Hospital Episode Statistics (HES) datasets. HES A&E collects data on all attendances to NHS EDs and includes basic information such as diagnosis, investigation, treatment, age, sex, area of residence and time and method of arrival and departure.

Emergency admissions to the same hospital were identified by linking the patient’s ED attendance with a discharge code of admission to HES APC emergency admissions in that same hospital using the pseudonymised patient identifier.

Table 1 shows how we selected the 13,912,890 ED attendances at major (type 1) NHS ED hospitals in 2018/19. These facilities are consultant-led, open 24 hours a day and offer full resuscitation facilities. The ‘minor’ types of ED include the other types of ED, such as type 2 EDs, which are for single specialities such as ophthalmology or dentistry and Type 3 and 4 EDs, such as minor injury units or NHS walk in centres, treat minor illnesses and conditions and may have limited opening hours and type 99 is attendance to an unknown type of ED unit. A previous study has shown that the distribution of attendances is uneven across England^15^.

**Table 1.** Selection of ED attendances for analysis.

|  |  |
| --- | --- |
| Number of all ED attendances | 22,367,847 |
| Drop duplicates | 22,289,194 |
| Keep only Type 1 ED attendances | 16,015,063 |
| Drop those who died in the department (154 NHS Trusts) | 16,001,011 |
| ED attendances merged with non-HES data (GP patient list and workforce, GPPS, QOF, the distance between LSOAs and ED of attendance and GP practice, rurality, GP extended access, QOF dimensions, Deprivation indices, ethnicity data from other HES attendances) | 15,995,619 |
| Excluding attendances from patients cared for by CCG's or GP's outside England | 15,469,684 |
| Excluding attendances at Specialist Trusts (women/children care Trusts) | 15,189,516 |
| Exclude attendances where GP practice has zero GPs | 15,084,005 |
| Excluding the three hospitals with high volume of direct admissions | 14,776,250 |
| Exclude ED attendances from patients which GP practice characteristics were missing | 13,912,890 |
| Exclude where there are other missing data. Final sample | 13,888,084 |
| Number of these ED attendances with an APC admission | 2,997,462 |
| Number of Clinical Commissioning Groups (CCGs) | 191 |
| Number of Hospital Trusts | 127 |

**Table 2.** Descriptive statistics of ED attendances.

| Variable | Mean | Std. Dev. |
| --- | --- | --- |
| Attendance characteristics (proportion) |  |  |
| Patient severity on attendance - included in 11 dummy variables according to treatment HRG | - | - |
| Week of year attended - included in 51 dummy variables | - | - |
| Emergency admission | 0.2 | 0.4 |
| Ambulance | 0.3 | 0.46 |
| Weekday | 0.72 | 0.45 |
| Weekday Core hours: 8 am-6:30 pm (GP open) | 0.44 | 0.5 |
| Week day AM extended hours: 6:30am-8 am (GP open) | 0 | 0.02 |
| Week day AM extended hours: 6:30am-8 am (GP closed) | 0 | 0.05 |
| Week day PM extended hours: 6:30pm-8pm (GP open) | 0.01 | 0.07 |
| Week day PM extended hours: 6:30pm-8pm (GP closed) | 0.01 | 0.09 |
| Weekend extended hours: 9:00 am-1pm (GP open) | 0 | 0.05 |
| Weekend extended hours: 9:00am - 1pm (GP closed) | 0.03 | 0.17 |
| Out of hours: Weekday (8pm-6:30am), Weekend (1pm<9am) | 0.41 | 0.49 |
| Patients demographics (Proportion) |  |  |
| Male | 0.49 | 0.5 |
| Age 0-4 | 0.1 | 0.3 |
| Age 5-17 | 0.12 | 0.33 |
| Age 18-65 | 0.53 | 0.5 |
| Age 65+ | 0.25 | 0.43 |
| Age included in 19 5-year bands as dummy variables | - | - |
| Patients residence characteristics (Proportion): |  |  |
| Urban | 0.87 | 0.34 |
| IMD quartile 1( <25th percentile)- Least deprived | 0.26 | 0.44 |
| IMD quartile 2 (25th-50th percentile) | 0.25 | 0.44 |
| IMD quartile 3 (50th-75th percentile) | 0.25 | 0.43 |
| IMD quartile 4 (>75th percentile)- Most deprived | 0.24 | 0.43 |
| Patients' GP practice characteristics: |  |  |
| Percentage of patients on GP practice disease registers - included in 21 variables | - | - |
| Percentage of patients aware that their GP practice has early morning extended hours | 10.57 | 9.47 |
| Percentage of patients aware that their GP practice has late afternoon extended hours | 12.77 | 8.59 |
| Percentage of patients aware that their GP practice has Saturday extended hours | 9.61 | 10.04 |
| Percentage of patients aware that they were able to see their preferred GP always or a lot | 45.39 | 16.87 |
| Percentage of patients aware that they were able to see their GP the same day | 32.86 | 13.51 |
| Percentage of patients aware that they were able to see their GP the next day | 10.99 | 5.66 |
| Percentage of patients that were very or fairly satisfied with the practice overall care | 83.08 | 9.57 |
| FTE GPs per 1000 patients | 0.55 | 0.23 |

We identify a number of attendance characteristics of the patients such as severity of the illness on attendance at the ED as proxied by the twelve ED Healthcare Resource Groups (HRGs), arrival by ambulance and non-urgent ED attendance (without the requirement for the patient not to be admitted following the attendance from the definition so this variable does not perfectly predict if the attendance resulted in an admission)^16^, along with patient demographic characteristics (e.g. gender, age band and ethnic group), patient’s area characteristics (such as their socio-economic status and urbanicity) and then link patients to their GP practice to provide further controls for case-mix (via the prevalence of illness in the practice), clinical quality and patient satisfaction with accessibility. For GP treatment quality, we included Quality Outcomes Framework (QOF) clinical points and General Practice Patient Survey measures on accessibility and satisfaction, focusing on awareness of extended hours and the ability to see a GP promptly. Workforce availability (GPs, nurses, staff per 1,000 patients) and morbidity indicators, like life expectancy and prevalence of key conditions, were also included, as was the distance from patients’ locations to their GP and nearest emergency department (ED). The availability of other ED services within 10km was also included to account for alternative emergency care options.

#### Analysis

Our goal is to establish variation between hospitals in the probability of a patient being admitted after visiting the ED, accounting for patient characteristics, their mode of attendance and the primary care setting that they come from. To achieve that we estimate a fixed effects regression model on individual-level ED attendance data. Our estimating equation is:

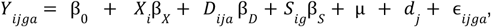

where *Y*_*ijga*_ is a binary indicator that takes the value 1 if there is an admission following ED attendance *a*, at hospital *j*, by patient *i* who is registered with GP practice *g*. The regression variables *X, D* and *S* are characteristics of the patient, of the attendance and of the GP practice of the specific patient *i*, respectively. The coefficients β_*X*_ β_*D*_ and β_*S*_ measure the effect of those variables on the probability of admission following the ED attendance. Central to our analysis is the fixed effect for the hospital, *d*_*j*_, which captures variation between hospitals that cannot be explained in terms of the foregoing characteristics. Finally ϵ_*jiga*_ is an idiosyncratic error term. We estimate the model using Stata® 17 with the package *xtreg* using the fixed effects option and specifying robust standard errors clustered at the hospital level. We use a linear probability model because our focus is on hospital fixed effects rather than forecasting individual patient predictors and this model is computationally efficient and avoids the incidental parameters problem^17^.

We estimate two further variants of the model above as robustness checks. In the first, we include additional fixed effects for the patient’s local healthcare authority, which at the time of the study was referred to as their Clinical Commissioning Group (CCG). This is to capture some further unobserved variation in patients’ access to emergency care which might not otherwise be captured and could also reflect otherwise unobserved elements of the severity of a patient’s illness. In the second variant, we estimate the above model, omitting the general practice characteristics as a check to ensure the results are not overly sensitive to these. The reason for this check is that we want to be sure that what we are establishing as variation in hospital decisions is not being driven by a patient’s primary care provision.

We use the model to calculate hospitals’ predicted admission rates defined as the percentage of patients attending a hospital’s ED that are expected to be admitted based on their characteristics.

## Results

### Descriptive statistics

Table 1 reports the descriptive statistics for attendance, patient and GP practice characteristics. 20% of ED attendances result in emergency admissions. 44% of ED attendances are during weekday core hours. 30% of patients arrive at the ED by ambulance. 49% of attendances are from male patients, 87% are from urban areas, 26% are from patients in the lowest deprivation quintile while 24% of patients are from the highest deprivation quintile. In terms of age, 10% of attendances are from patients in the age group 0-4 years, and 25% are from patients aged 65 and above.

Patients are generally satisfied with the GP practices overall care (83%) and their ability to see their preferred GP (45%) but are unlikely to know about the practices arrangement for extended hours services.

Figure 1 shows a histogram of ED admission rates across hospitals.

**Figure 1.**
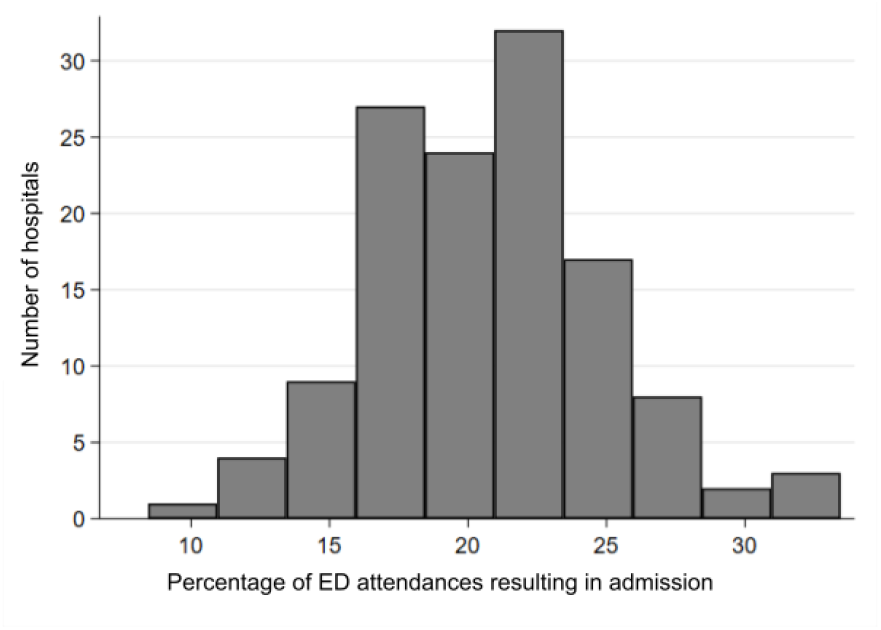
The distribution of admission rates across hospitals

To illustrate variation in ED admission across hospitals, figure 2 provides a caterpillar plot of admission rates for all 127 hospitals in our data before making any adjustments for differing patient characteristics. Hospitals are ranked on the horizontal axis from lowest to highest admission rates. The lowest admitting hospitals have rates of less than 15%, whilst there are approximately 20 hospitals with a rate greater than 25%.

**Figure 2.**
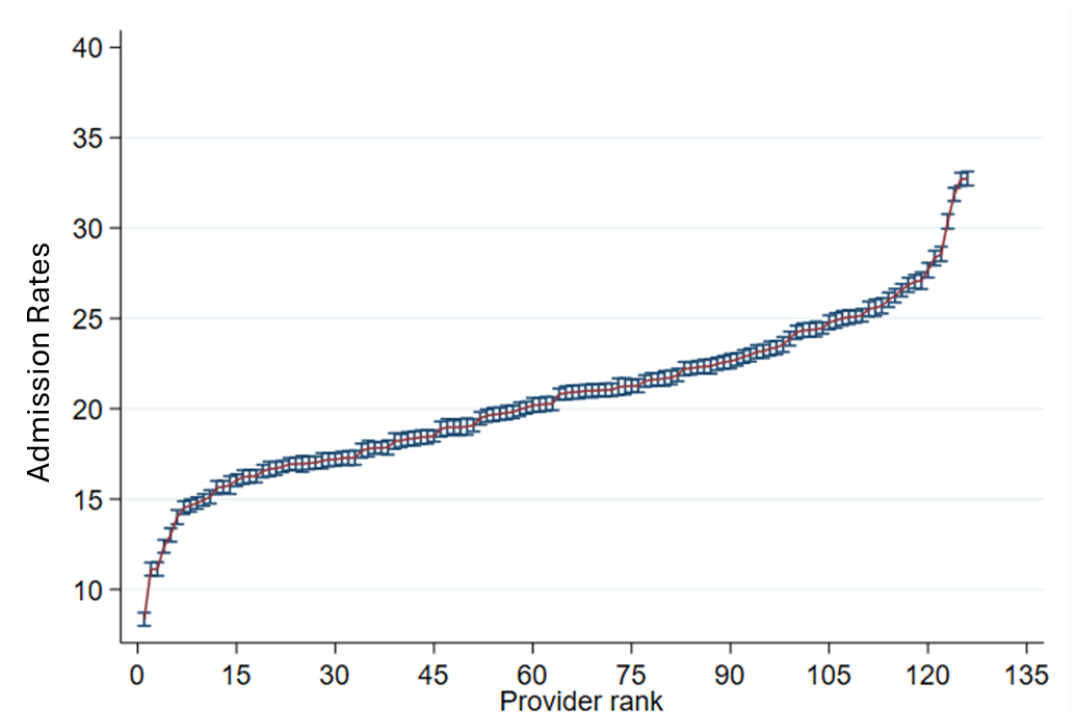
Hospital admission rates without adjusting for case-mix

Since each observation in figure 2 relates to a specific hospital, their locations can be plotted, and the associated admission rate can then be illustrated for the areas they serve, as shown in figure 3.

**Figure 3.**
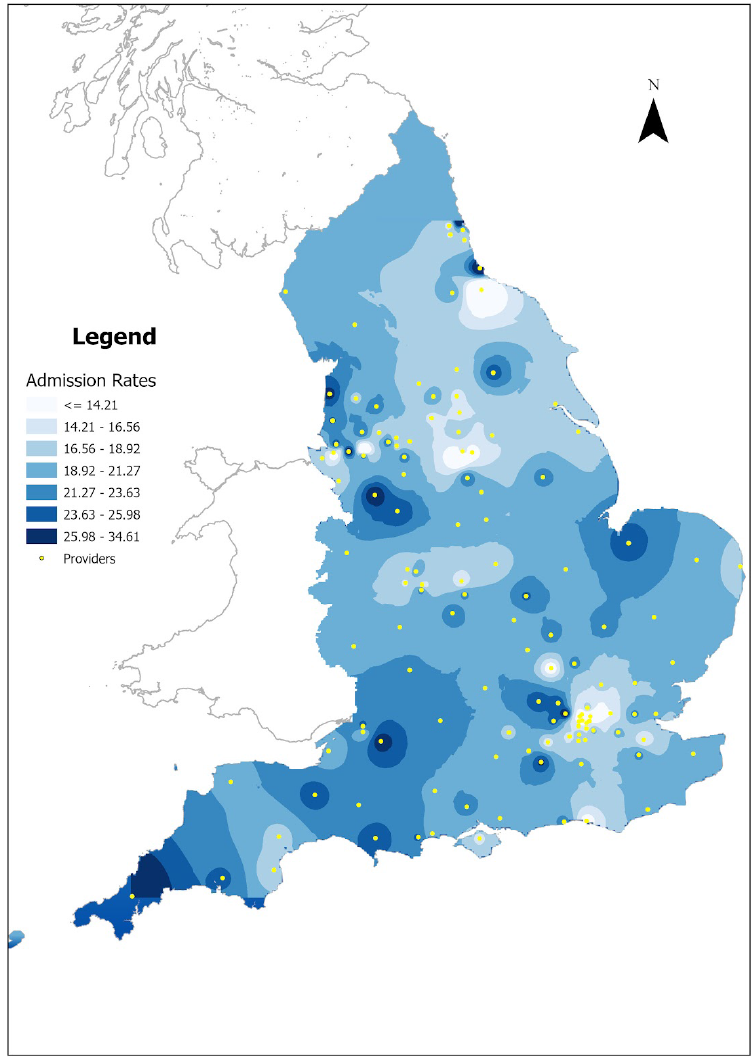
Spatial distribution of hospitals’ admission rates without case-mix adjustment

The darker regions in the figure correspond to areas where hospitals with higher admission rates are clustered. There is an uneven distribution of these higher admission areas, focused on the far west, the north west and the southern midlands in England. This is useful contextual information for the purposes of policy because the NHS in England is organised on a regional basis and the figure shows those regions which potentially have the greatest problem in terms of ED admissions. However, the figure is drawn without taking into account the different characteristics of patients who attend these different hospitals, so we turn next to our regression model.

### Model results

Our model estimated the probability of a patient being admitted and included a total of 153 variables describing the patient’s characteristics, the characteristics of the local area they reside in, the nature of their attendance, and the characteristics of the GP practice responsible for their primary care. Three versions of the model were estimated as described above, and the full regression results table for all three versions is available in the online supplementary table. Across the three models, there is significant variation in hospital admission probabilities after ED attendance, influenced by patient demographics, socioeconomic status, and accessibility. Older adults, non-White ethnicities, and residents of deprived areas had higher admission probabilities. Accessibility factors, like distance to the ED, ambulance arrival and arrivals during closed GP hours, increased admission likelihood. Perhaps surprisingly, patients who live nearer their GP are more likely to be admitted when they attend ED, which could indicate that close access to a GP provides emergency care for less severe patients, so that only the more severe attend an ED. GP practice characteristics, including awareness of extended hours, staff levels, and prevalence of chronic diseases also impacted ED attendance. Adjustments for CCG and hospital-specific factors highlighted unobserved variations, consistent with the baseline model results. Overall, the models account for between 15% and 17% of the variation in admission. A Hausman test of random effects rejects the null hypothesis that the errors and regressors are not correlated, and hence validates the use of the fixed effects specification.

The regression models can be used to construct analogues of figures 2 and 3, taking into account all the confounding variables. figure 4 is a caterpillar plot of the variation of *adjusted percentages* of admissions from our baseline model estimates. Whilst the identities of the individual hospitals differ from those in figure 2, the overall pattern and degree of dispersion in admission rates are very similar. Hence, adjusting for hospitals’ case mix in respect of patient and attendance characteristics reorders some hospitals but does not reduce overall variation to any great extent. There remain hospitals with admission rates of less than 15%, while there are approximately 12 hospitals with a rate greater than 25%.

**Figure 4.**
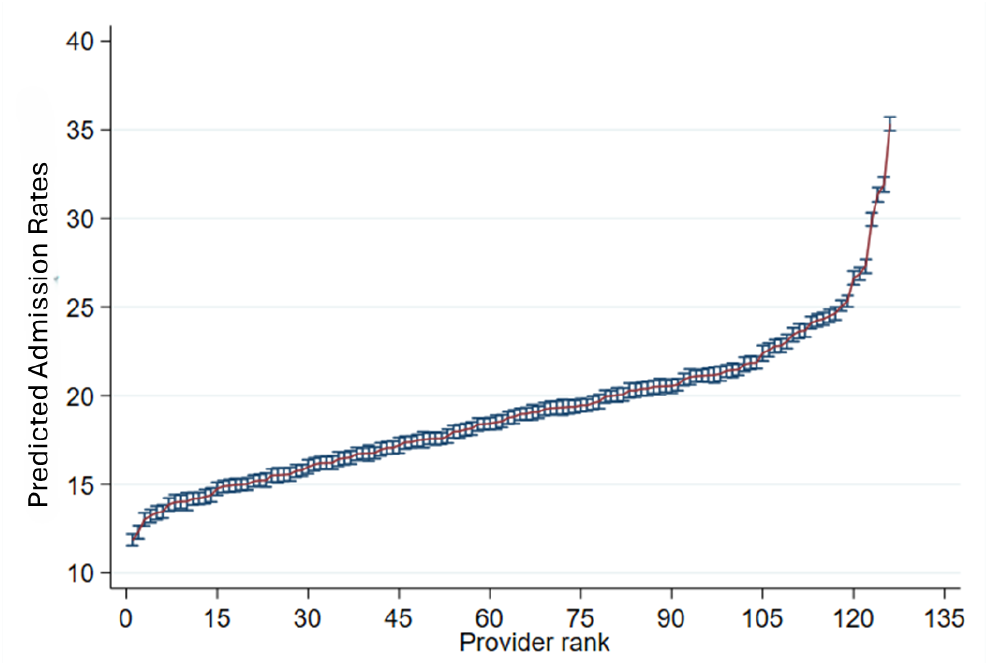
Predicted hospital admission rates after adjusting for case-mix

The results are similar across all three sets of analyses indicating that our results are robust. The relationship between unadjusted and adjusted admission rates for individual hospitals is shown in figure 5.

**Figure 5.**
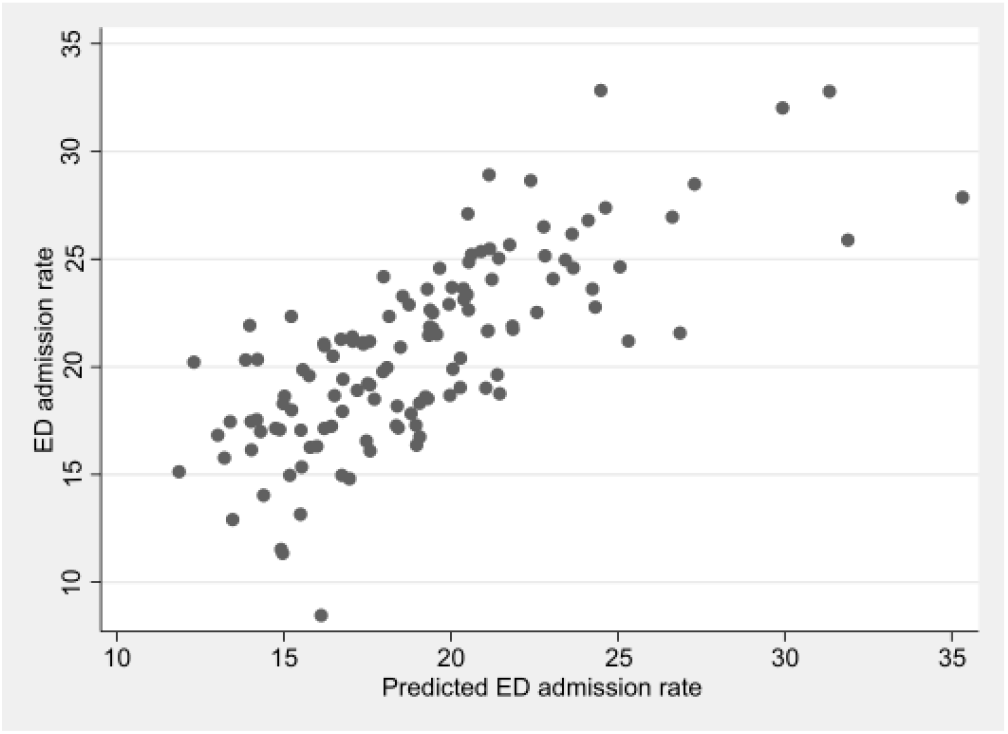
Relationship between unadjusted and adjusted ED admission rates

The spatial distribution of adjusted admission rates across hospitals is shown in figure 6. It shows fewer areas in darkest blue, i.e. with a higher percentage of emergency admissions following an ED attendance. The Liverpool, Bradford, Buckinghamshire and Guildford areas still have the highest admission rates. Whilst, Newcastle, York, Coventry and Norfolk have admission rates roughly close to the average, after adjustment. There are more broadly dispersed areas which show above average admission rates, covering much of the south, and an area from the north west to the north east extending into the midlands. In this adjusted version Yorkshire and the Humber are notable for having low admission rates indicating that the higher rates depicted in figure 4 can be explained by patient and attendance characteristics.

**Figure 6.**
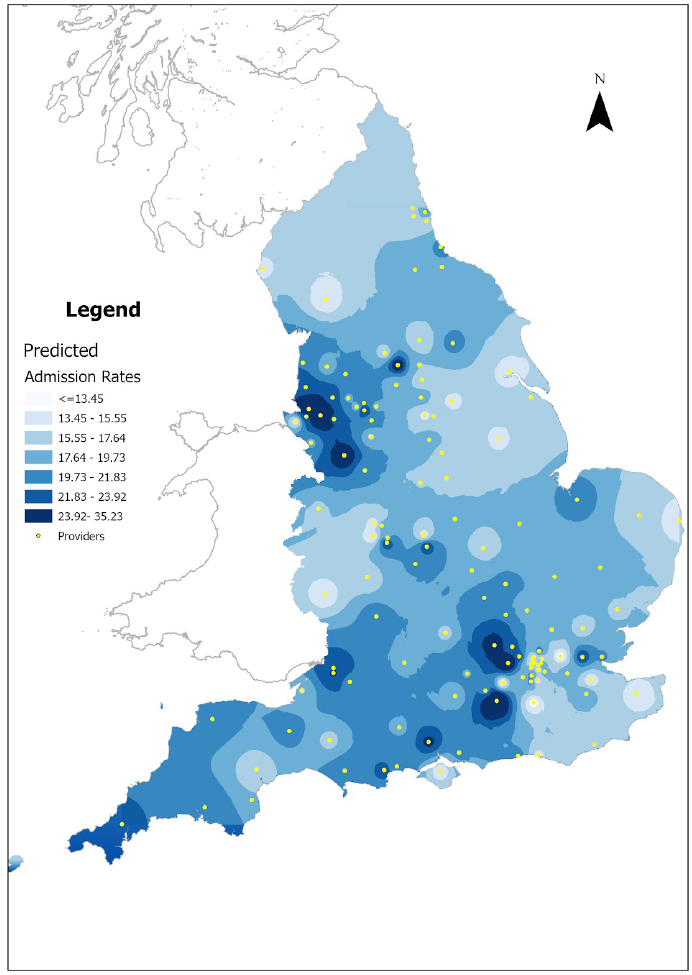
Spatial distribution of hospitals’ adjusted admission rates

## Discussion

### Key Findings

Our analysis finds that there is substantial and persistent variability in hospital admission rates following an emergency attendance across English NHS hospitals, even after controlling for a comprehensive range of patient and admission characteristics. On average, 20% of ED attendances resulted in emergency admissions. Explaining these differences we find that patient characteristics influencing admission probability include: a higher likelihood for older adults, non-White ethnic groups and patients from more deprived areas. Increased likelihood of admissions for patients arriving by an ambulance or during periods when GP practices were closed and for patients living closer to their GP. GP practice characteristics, such as awareness of extended hours, staff levels, and chronic disease prevalence also impacted admission outcomes. Even after extensive controls, variation in admission probabilities persisted among hospitals, ranging from 12% to 35%. The spatial distribution of higher adjusted admission rates is uneven, with areas like Liverpool, Bradford, Buckinghamshire, and Guildford still showing the highest rates. In contrast, areas like Newcastle, York, Coventry, and Norfolk have around average rates after adjustment.

## Limitations

As with any study based on administrative data, there is susceptibility to issues of completeness, accuracy and coding.

Our approach to identifying the residual variation in ED admission rates across hospitals is predicated on having good measures of the severity of a patient’s illness. We proxy this primarily using the twelve ED HRGs. HRGs are broad categories, and while useful, they do not fully capture clinical severity. This means that some unobserved patient-level severity factors will contribute to the unexplained variation in admission rates. To counter this we include extensive controls (153 variables), but unobserved factors such as differences in hospital culture, clinical thresholds for admission, bed availability or diagnostic capabilities are not measured or observed in the data.

The study uses data for 2018/19. Healthcare systems, policies, and patient behaviours evolve over time (e.g., due to the COVID-19 pandemic or subsequent NHS reforms). The findings, while robust for the period, might not reflect the current state of emergency admissions in 2025.

## Implications for policy

A key policy implication is that a uniform national approach to managing emergency admissions is not likely to be effective. Hence, policymakers should avoid ‘one-size-fits-all’ directives for ED admissions, as these fail to address the underlying local and hospital-specific factors driving the observed variations.This study contributes important evidence for policy, indicating that a uniform approach to managing admissions may overlook critical regional and demographic factors. A tailored strategy, which is directed at local resource availability, patient needs and hospital capabilities may be required.

The importance in our regression results of proximity to other facilities suggests that well-placed urgent care facilities and improved transport options could alleviate pressure on emergency services in certain areas.The findings that older adults, non-White ethnic groups, and patients from more deprived areas have a higher likelihood of admission point to the need for policies that address health inequalities. Furthermore, the finding that patients living closer to their GP are more likely to be admitted having attended an ED warrants further investigation from a policy perspective, as it might indicate that proximity to a GP acts as a filter keeping less seriously ill patients out of an ED.

Our approach of linking ED attendances with GP practice characteristics also provides a possible avenue for policy development. Future policy might usefully exploit similar integrated data to provide insights into patient pathways and systemic factors that contribute to admissions.

Future research should aim to integrate these dimensions further, exploring the role of alternative emergency care services, patient socioeconomic factors, and broader emergency care infrastructure to build a more responsive and equitable health system across regions.

## Supporting information

table of full regression results

## Data Availability

All data used are available from NHS England Digital upon registration of an appropriate data sharing agreement.

https://digital.nhs.uk/services/hospital-episode-statistics

## Supplementary material only to be be made available online

### S1 Sample selection notes

We excluded the following:

1. Type 2, 3, 4 and type 99 ED attendances and only use Type 1 as these are consultant led, open 24 hours per day and have full resuscitation facilities. Type 2 EDs are for single specialities such as ophthalmology or dentistry and Type 3 and 4 EDs, such as minor injury units or NHS walk in centres, treat minor illnesses and conditions and may have limited opening hours and type 99 is attendance to an unknown type of ED unit. This was because the provision of ‘minor’ ED services varies across the country and these services cater to a patient population that is typically not at risk of admission to inpatient care, that is, a population that is not the focus of this analysis.
2. Those that died in the department and those that were admitted via non-ED admission method as this is not the population of interest for our analysis.
3. Trusts with 0, 5 and N. We excluded primary care trusts, trusts with associated treatment centres, NHS trust treatment centres listed separately to NHS trusts, independent hospitals and other independent sector healthcare providers.
4. Exclusion of direct admissions. We dropped three hospitals RC9, RA9, RTR that had direct admissions.
5. Commissioning hubs and specialist trusts that care for women and children. We exclude the 30 commissioning hubs (NHS Digital) from the analysis since they are responsible for leading the commissioning of specialised services for a wider population.
6. Patients cared for by CCGs or GPs outside England. We excluded ED attendances by people from other parts of the UK, or attendances of English patients outside England and attendances to ED units run by independent providers because these attendances would be covered by separate contracts with CCGs.
7. Using CCG codes from the QOF 2018-19 report, we found that some CCGs had merged (On April 1, 2019 NHS Erewash, Hardwick, North Derbyshire and South Derbyshire CCGs merged to form NHS Derbyshire. NHS North, East, West Devon CCG and NHS South Devon and Torbay merged into NHS Devon). According to the “CCG level exceptions and exclusions report table” there were 191 CCGs in 2018/19.
8. We found that there were 194 CCGs in October 2018 and in March 2019. However, in March 2020 there were 191 CCGs. So, we report 191 CCGs for the analysis.
9. After eliminating observations that fall in one or more of these categories, the final estimation sample consists of 13,912,890 ED attendances with 2,997,462 admissions across 127 trusts and 191 CCGs for the period 2018-19.
10. The non-missing data for regression analysis consists of 13,888,084 ED attendances with 2,835,283 admissions.

### S2 Full regression analysis

[Full regression results](https://docs.google.com/spreadsheets/d/1uvcb67geWjxUliaXA867Xy9eYM5XkluEWQZ3aOiekxQ/edit?gid=0#gid=0)

