## Supplementary material for "Variation in admissions from hospital emergency departments in the English NHS": table of full regression results

**Regression estimates for probability of admission, hospital fixed effect**

Coeffients with standard errors in the line below

| | Baseline | With CCG fixed effects | Without GP practice characteristics |
|---|---|---|---|
| *Patient characteristics* | | | |
| Age 6-10 (under 6 years as the reference group) | -6.305*** | -6.288*** | -6.308*** |
| | -0.057 | -0.057 | -0.056 |
| Age 11-15 (under 6 years as the reference group) | -7.850*** | -7.834*** | -7.826*** |
| | -0.055 | -0.055 | -0.054 |
| Age 16-20 (under 6 years as the reference group) | -7.808*** | -7.834*** | -7.839*** |
| | -0.052 | -0.052 | -0.051 |
| Age 21-25 (under 6 years as the reference group) | -7.441*** | -7.455*** | -7.498*** |
| | -0.05 | -0.05 | -0.049 |
| Age 26-30 (under 6 years as the reference group) | -6.660*** | -6.666*** | -6.707*** |
| | -0.049 | -0.049 | -0.048 |
| Age 31-35 (under 6 years as the reference group) | -6.013*** | -6.021*** | -6.067*** |
| | -0.051 | -0.051 | -0.05 |
| Age 36-40 (under 6 years as the reference group) | -5.450*** | -5.463*** | -5.494*** |
| | -0.053 | -0.053 | -0.052 |
| Age 41-45 (under 6 years as the reference group) | -4.900*** | -4.916*** | -4.970*** |
| | -0.055 | -0.055 | -0.054 |
| Age 46-50 (under 6 years as the reference group) | -3.848*** | -3.861*** | -3.871*** |
| | -0.054 | -0.054 | -0.053 |
| Age 51-55 (under 6 years as the reference group) | -2.837*** | -2.848*** | -2.851*** |
| | -0.054 | -0.054 | -0.053 |
| Age 56-60 (under 6 years as the reference group) | -1.235*** | -1.248*** | -1.234*** |
| | -0.056 | -0.056 | -0.055 |
| Age 61-65 (under 6 years as the reference group) | 0.676*** | 0.666*** | 0.720*** |
| | -0.059 | -0.059 | -0.058 |
| Age 66-70 (under 6 years as the reference group) | 2.526*** | 2.512*** | 2.582*** |

| | | | |
|---|---|---|---|
| | -0.059 | -0.059 | -0.058 |
| Age 71-75 (under 6 years as the reference group) | 4.056*** | 4.049*** | 4.106*** |
| | -0.057 | -0.057 | -0.056 |
| Age 76-80 (under 6 years as the reference group) | 5.184*** | 5.186*** | 5.240*** |
| | -0.058 | -0.058 | -0.057 |
| Age 81-85 (under 6 years as the reference group) | 6.142*** | 6.151*** | 6.188*** |
| | -0.059 | -0.059 | -0.058 |
| Age 86-90 (under 6 years as the reference group) | 6.849*** | 6.860*** | 6.903*** |
| | -0.064 | -0.064 | -0.063 |
| Age 91-95 (under 6 years as the reference group) | 8.115*** | 8.121*** | 8.214*** |
| | -0.084 | -0.084 | -0.082 |
| Age 96+ (under 6 years as the reference group) | 9.453*** | 9.470*** | 9.506*** |
| | -0.148 | -0.147 | -0.145 |
| Male gender | -0.479*** | -0.486*** | -0.486*** |
| | -0.02 | -0.02 | -0.02 |
| Mixed ethnicity groups (Ref: White Ethnicity) | 0.570*** | 0.596*** | 0.471*** |
| | -0.074 | -0.074 | -0.072 |
| Asian ethnicity (Ref: White Ethnicity) | 1.018*** | 1.074*** | 0.894*** |
| | -0.043 | -0.043 | -0.039 |
| Black ethnicity (Ref: White Ethnicity) | 0.730*** | 0.810*** | 0.596*** |
| | -0.053 | -0.054 | -0.052 |
| Other ethnic group (Ref: White Ethnicity) | 0.348*** | 0.409*** | 0.210*** |
| | -0.061 | -0.061 | -0.059 |
| Unknown ethnicity (Ref: White Ethnicity) | -0.065 | -0.099* | -0.131** |
| | -0.058 | -0.058 | -0.057 |
| *Patient's LSOA characteristics* | | | |
| Living in third most deprived quartile of LSOAs (Ref: least deprived LSOA quartile) | -0.091*** | -0.099*** | -0.121*** |
| | -0.029 | -0.029 | -0.028 |
| Living in second most deprived quartile of LSOAs (Ref: least deprived LSOA quartile) | -0.312*** | -0.319*** | -0.390*** |

| | | | |
|---|---|---|---|
| | -0.032 | -0.032 | -0.029 |
| Living in most deprived quartile of LSOAs (Ref: least deprived LSOA quartile) | -0.689*** | -0.691*** | -0.815*** |
| | -0.036 | -0.037 | -0.032 |
| Distance to AED of attendance between 10 and 20 km (Ref: Distance to AED of attendance of less than 10km) | 1.107*** | 1.056*** | 1.215*** |
| | -0.039 | -0.041 | -0.038 |
| Distance to AED of attendance more than 20km (Ref: Distance to AED of attendance less than 10km) | 1.210*** | 1.206*** | 1.352*** |
| | -0.043 | -0.046 | -0.042 |
| Distance to GP practice between 1 and 2 km (Ref: Distance to GP practice is less than 1km) | 0.069*** | 0.011 | 0.083*** |
| | -0.024 | -0.024 | -0.024 |
| Distance to GP practice of more than 2km (Ref: Distance to GP practice is less than 1km) | -0.114*** | -0.160*** | -0.150*** |
| | -0.026 | -0.026 | -0.025 |
| Patient lives in urban area | -0.205*** | -0.228*** | -0.443*** |
| | -0.037 | -0.038 | -0.034 |
| *Attendance characteristics* | | | |
| Arriving by ambulance | 10.821*** | 10.765*** | 10.812*** |
| | -0.025 | -0.025 | -0.025 |
| Arriving during weekday AM extended hours w/ GP open (Ref: Arriving during core GP hours) | 0.112 | 0.094 | 0.14 |
| | -0.427 | -0.426 | -0.42 |
| Arriving during weekday AM extended hours w/ GP closed (Ref: Arriving during core GP hours) | 0.997*** | 0.977*** | 0.991*** |
| | -0.194 | -0.194 | -0.19 |
| Arriving during weekday PM extended hours w/ GP open (Ref: Arriving during core GP hours) | -1.983*** | -2.010*** | -1.992*** |
| | -0.149 | -0.149 | -0.147 |
| Arriving during weekday PM extended hours w/ GP closed (Ref: Arriving during core GP hours) | -1.886*** | -1.886*** | -1.843*** |
| | -0.121 | -0.121 | -0.118 |
| Arriving during weekend extended hours w/ GP open (Ref: Arriving during core GP hours) | 0.066 | 0.054 | -0.009 |

| | | | |
|---|---|---|---|
| | -0.221 | -0.222 | -0.217 |
| Arriving during weekend extended hours w/ GP closed (Ref: Arriving during core GP hours) | -0.358*** | -0.360*** | -0.350*** |
| | -0.068 | -0.068 | -0.067 |
| Arriving out of all GP hours (Ref: Arriving during core GP hours) | -0.993*** | -1.009*** | -0.983*** |
| | -0.025 | -0.025 | -0.024 |
| HRG UZ01Z, Invalid grouping (Ref: HRG VB11Z, No Investigation with No Significant Treatment | 25.914*** | 25.981*** | 25.831*** |
| | -1.518 | -1.518 | -1.482 |
| HRG VB01Z, Any Investigation with Category 5 Treatment (Ref: HRG VB11Z, No Investigation with No Significant Treatment) | 39.444*** | 39.348*** | 39.278*** |
| | -1.4 | -1.399 | -1.363 |
| HRG VB02Z, Category 3 Investigation with Category 4 Treatment (Ref: HRG VB11Z, No Investigation with No Significant Treatment) | 39.508*** | 39.510*** | 39.425*** |
| | -1.376 | -1.375 | -1.34 |
| HRG VB03Z, Category 3 Investigation with Category 1-3 Treatment (Ref: HRG VB11Z, No Investigation with No Significant Treatment) | 25.624*** | 25.627*** | 25.483*** |
| | -1.374 | -1.374 | -1.338 |
| HRG VB04Z, Category 2 Investigation with Category 4 Treatment (Ref: HRG VB11Z, No Investigation with No Significant Treatment) | 34.843*** | 34.893*** | 34.720*** |
| | -1.374 | -1.374 | -1.338 |
| HRG VB05Z, Category 2 Investigation with Category 3 Treatment (Ref: HRG VB11Z, No Investigation with No Significant Treatment) | 17.344*** | 17.392*** | 17.241*** |
| | -1.376 | -1.375 | -1.34 |
| HRG VB06Z, Category 1 Investigation with Category 3-4 Treatment (Ref: HRG VB11Z, No Investigation with No Significant Treatment) | 11.427*** | 11.473*** | 11.362*** |
| | -1.375 | -1.375 | -1.339 |
| HRG VB07Z, Category 2 Investigation with Category 2 Treatment (Ref: HRG VB11Z, No Investigation with No Significant Treatment) | 13.285*** | 13.342*** | 13.179*** |
| | -1.374 | -1.374 | -1.338 |
| HRG VB08Z, Category 2 Investigation with Category 1 Treatment (Ref: HRG VB11Z, No Investigation with No Significant Treatment) | 10.961*** | 11.004*** | 10.809*** |
| | -1.374 | -1.374 | -1.338 |
| HRG VB09Z, Category 1 Investigation with Category 1-2 Treatment (Ref: HRG VB11Z, No Investigation with No Significant Treatment) | 3.315** | 3.370** | 3.149** |

| | | | |
|---|---|---|---|
| | -1.374 | -1.373 | -1.338 |
| HRG VB10Z, Dental Care (Ref: HRG VB11Z, No Investigation with No Significant Treatment) | 3.450** | 3.485** | 3.256** |
| | -1.374 | -1.373 | -1.338 |
| Nearest AED provider to patient's LSOA is a type1 (Ref: nearest AED provider to patient's LSOA is a type 4) | -0.606*** | -0.407*** | -0.571*** |
| | -0.052 | -0.055 | -0.051 |
| Nearest AED provider to patient's LSOA is a type2 (Ref: nearest AED provider to patient's LSOA is a type 4) | -0.581*** | -0.430*** | -0.606*** |
| | -0.066 | -0.068 | -0.064 |
| Nearest AED provider to patient's LSOA is a type3 (Ref: nearest AED provider to patient's LSOA is a type 4) | -0.235*** | -0.177*** | -0.195*** |
| | -0.054 | -0.056 | -0.052 |
| Nearest AED provider to patient's LSOA is a type99 (Ref: nearest AED provider to patient's LSOA is a type 4) | -0.165 | 0.109 | -0.15 |
| | -0.145 | -0.147 | -0.138 |
| Patient's LSOA is within a 10km radius of a type1 AED | -0.897*** | -0.929*** | -0.950*** |
| | -0.044 | -0.048 | -0.043 |
| Patient's LSOA is within a 10km radius of a type2 AED | -0.816*** | -0.334*** | -1.103*** |
| | -0.039 | -0.045 | -0.037 |
| Patient's LSOA is within a 10km radius of a type3 AED | 0.550*** | 0.554*** | 0.502*** |
| | -0.032 | -0.036 | -0.032 |
| Patient's LSOA is within a 10km radius of a type4 AED | 0.249*** | 0.109** | 0.356*** |
| | -0.041 | -0.049 | -0.04 |
| Patient's LSOA is within a 10km radius of a type99 AED | -0.275*** | 0.384*** | -0.366*** |
| | -0.085 | -0.097 | -0.082 |
| Avoidable admission (using modified NHS digital admission) | -2.889*** | -2.841*** | -2.847*** |
| | -0.043 | -0.043 | -0.042 |
| Monday (Ref:Friday) | -0.769*** | -0.766*** | -0.755*** |
| | -0.038 | -0.038 | -0.037 |
| Saturday (Ref:Friday) | -1.118*** | -1.105*** | -1.110*** |
| | -0.041 | -0.041 | -0.04 |
| Sunday (Ref:Friday) | -1.428*** | -1.414*** | -1.416*** |

| | | | |
|---|---|---|---|
| | -0.045 | -0.045 | -0.044 |
| Thursday (Ref: Friday) | -0.292*** | -0.291*** | -0.300*** |
| | -0.039 | -0.039 | -0.038 |
| Tuesday (Ref: Friday) | -0.615*** | -0.614*** | -0.611*** |
| | -0.039 | -0.039 | -0.038 |
| Wednesday (Ref: Friday) | -0.428*** | -0.426*** | -0.432*** |
| | -0.039 | -0.039 | -0.038 |
| 2018 , week 14 (Ref: 2018, week 13) | -0.803*** | -0.801*** | -0.773*** |
| | -0.21 | -0.21 | -0.206 |
| 2018 , week 15 (Ref: 2018, week 13) | -0.594*** | -0.592*** | -0.589*** |
| | -0.21 | -0.21 | -0.206 |
| 2018 , week 16 (Ref: 2018, week 13) | -0.635*** | -0.630*** | -0.629*** |
| | -0.21 | -0.21 | -0.205 |
| 2018 , week 17 (Ref: 2018, week 13) | -0.305 | -0.304 | -0.293 |
| | -0.21 | -0.21 | -0.205 |
| 2018 , week 18 (Ref: 2018, week 13) | -0.418** | -0.414** | -0.404** |
| | -0.209 | -0.209 | -0.205 |
| 2018 , week 19 (Ref: 2018, week 13) | -1.237*** | -1.233*** | -1.221*** |
| | -0.209 | -0.209 | -0.205 |
| 2018 , week 20 (Ref: 2018, week 13) | -1.104*** | -1.097*** | -1.093*** |
| | -0.209 | -0.209 | -0.205 |
| 2018 , week 21 (Ref: 2018, week 13) | -0.906*** | -0.908*** | -0.884*** |
| | -0.209 | -0.209 | -0.205 |
| 2018 , week 22 (Ref: 2018, week 13) | -1.184*** | -1.177*** | -1.204*** |
| | -0.209 | -0.209 | -0.205 |
| 2018 , week 23 (Ref: 2018, week 13) | -0.956*** | -0.954*** | -0.925*** |
| | -0.209 | -0.209 | -0.205 |
| 2018 , week 24 (Ref: 2018, week 13) | -1.026*** | -1.025*** | -1.040*** |
| | -0.209 | -0.209 | -0.205 |
| 2018 , week 25 (Ref: 2018, week 13) | -0.921*** | -0.922*** | -0.938*** |

| | | | |
|---|---|---|---|
| | -0.209 | -0.209 | -0.205 |
| 2018 , week 26 (Ref: 2018, week 13) | -1.335*** | -1.328*** | -1.337*** |
| | -0.209 | -0.209 | -0.205 |
| 2018 , week 27 (Ref: 2018, week 13) | -1.581*** | -1.574*** | -1.565*** |
| | -0.209 | -0.209 | -0.204 |
| 2018 , week 28 (Ref: 2018, week 13) | -1.676*** | -1.670*** | -1.685*** |
| | -0.209 | -0.209 | -0.205 |
| 2018 , week 29 (Ref: 2018, week 13) | -1.441*** | -1.436*** | -1.443*** |
| | -0.209 | -0.209 | -0.205 |
| 2018 , week 30 (Ref: 2018, week 13) | -1.582*** | -1.583*** | -1.577*** |
| | -0.21 | -0.21 | -0.205 |
| 2018 , week 31 (Ref: 2018, week 13) | -1.381*** | -1.378*** | -1.365*** |
| | -0.21 | -0.21 | -0.206 |
| 2018 , week 32 (Ref: 2018, week 13) | -1.345*** | -1.342*** | -1.324*** |
| | -0.21 | -0.21 | -0.206 |
| 2018 , week 33 (Ref: 2018, week 13) | -1.052*** | -1.048*** | -1.037*** |
| | -0.21 | -0.21 | -0.206 |
| 2018 , week 34 (Ref: 2018, week 13) | -1.250*** | -1.248*** | -1.240*** |
| | -0.21 | -0.21 | -0.206 |
| 2018 , week 35 (Ref: 2018, week 13) | -1.319*** | -1.312*** | -1.293*** |
| | -0.21 | -0.21 | -0.206 |
| 2018 , week 36 (Ref: 2018, week 13) | -1.202*** | -1.199*** | -1.182*** |
| | -0.21 | -0.21 | -0.206 |
| 2018 , week 37 (Ref: 2018, week 13) | -1.041*** | -1.035*** | -1.037*** |
| | -0.209 | -0.209 | -0.205 |
| 2018 , week 38 (Ref: 2018, week 13) | -1.035*** | -1.031*** | -1.031*** |
| | -0.209 | -0.209 | -0.205 |
| 2018 , week 39 (Ref: 2018, week 13) | -1.039*** | -1.032*** | -1.030*** |
| | -0.209 | -0.209 | -0.205 |
| 2018 , week 40 (Ref: 2018, week 13) | -1.066*** | -1.064*** | -1.082*** |

| | | | |
|---|---|---|---|
| | -0.209 | -0.209 | -0.205 |
| 2018 , week 41 (Ref: 2018, week 13) | -0.888*** | -0.888*** | -0.879*** |
| | -0.209 | -0.209 | -0.205 |
| 2018 , week 42 (Ref: 2018, week 13) | -0.909*** | -0.904*** | -0.907*** |
| | -0.209 | -0.209 | -0.205 |
| 2018 , week 43 (Ref: 2018, week 13) | -0.534** | -0.538** | -0.547*** |
| | -0.21 | -0.21 | -0.206 |
| 2018 , week 44 (Ref: 2018, week 13) | -0.757*** | -0.758*** | -0.728*** |
| | -0.21 | -0.21 | -0.205 |
| 2018 , week 45 (Ref: 2018, week 13) | -0.620*** | -0.621*** | -0.625*** |
| | -0.209 | -0.209 | -0.205 |
| 2018 , week 46 (Ref: 2018, week 13) | -0.724*** | -0.724*** | -0.722*** |
| | -0.209 | -0.209 | -0.205 |
| 2018 , week 47 (Ref: 2018, week 13) | -0.356* | -0.355* | -0.384* |
| | -0.209 | -0.209 | -0.205 |
| 2018 , week 48 (Ref: 2018, week 13) | -0.567*** | -0.569*** | -0.593*** |
| | -0.209 | -0.209 | -0.205 |
| 2018 , week 49 (Ref: 2018, week 13) | -1.014*** | -1.013*** | -1.005*** |
| | -0.209 | -0.209 | -0.205 |
| 2018 , week 50 (Ref: 2018, week 13) | -0.685*** | -0.683*** | -0.679*** |
| | -0.209 | -0.209 | -0.205 |
| 2018 , week 51 (Ref: 2018, week 13) | -0.552*** | -0.544*** | -0.575*** |
| | -0.209 | -0.209 | -0.205 |
| 2018 , week 52 (Ref: 2018, week 13) | -0.274 | -0.272 | -0.281 |
| | -0.209 | -0.208 | -0.204 |
| 2019, week 1 (Ref: 2018, week 13) | -1.096*** | -1.094*** | -1.082*** |
| | -0.209 | -0.209 | -0.205 |
| 2019, week 2 (Ref: 2018, week 13) | -0.905*** | -0.904*** | -0.897*** |
| | -0.209 | -0.209 | -0.205 |
| 2019, week 3 (Ref: 2018, week 13) | -0.711*** | -0.707*** | -0.711*** |

| | | | |
|---|---|---|---|
| | -0.209 | -0.209 | -0.205 |
| 2019, week 4 (Ref: 2018, week 13) | -0.712*** | -0.710*** | -0.687*** |
| | -0.209 | -0.209 | -0.205 |
| 2019, week 5 (Ref: 2018, week 13) | -0.894*** | -0.892*** | -0.897*** |
| | -0.209 | -0.209 | -0.205 |
| 2019, week 6 (Ref: 2018, week 13) | -1.058*** | -1.053*** | -1.061*** |
| | -0.209 | -0.208 | -0.204 |
| 2019, week 7 (Ref: 2018, week 13) | -1.134*** | -1.129*** | -1.105*** |
| | -0.209 | -0.209 | -0.205 |
| 2019, week 8 (Ref: 2018, week 13) | -0.854*** | -0.850*** | -0.862*** |
| | -0.209 | -0.209 | -0.205 |
| 2019, week 9 (Ref: 2018, week 13) | -0.732*** | -0.730*** | -0.728*** |
| | -0.209 | -0.209 | -0.205 |
| 2019, week 10 (Ref: 2018, week 13) | -0.559*** | -0.556*** | -0.579*** |
| | -0.209 | -0.209 | -0.205 |
| 2019, week 11 (Ref: 2018, week 13) | -0.638*** | -0.635*** | -0.619*** |
| | -0.209 | -0.209 | -0.205 |
| 2019, week 12 (Ref: 2018, week 13) | -1.277*** | -1.272*** | -1.283*** |
| | -0.209 | -0.209 | -0.205 |
| 2019, week 13 (Ref: 2018, week 13) | -6.552*** | -6.547*** | -6.516*** |
| | -0.211 | -0.211 | -0.207 |
| *GP practice characteristics* | | | |
| % clinical QOF points in 2018 | -0.002 | 0.008*** | |
| | -0.002 | -0.002 | |
| % patients aware that GP has AM extended hrs | -0.007*** | -0.003*** | |
| | -0.001 | -0.001 | |
| % patients aware that GP has PM extended hrs | -0.002* | -0.001 | |
| | -0.001 | -0.001 | |
| % patients aware that GP has Sat extended hrs | -0.001 | 0 | |
| | -0.001 | -0.001 | |

| | | |
|---|---|---|
| % patients able to see pref GP (always or a lot) | 0.001* | 0.001 |
| | -0.001 | -0.001 |
| % patients able to see GP the same day | 0.003*** | 0 |
| | -0.001 | -0.001 |
| % patients able to see GP the next day | 0.002 | 0.004** |
| | -0.002 | -0.002 |
| % patients very or fairly satisfied with GP care | 0.003** | 0.004*** |
| | -0.002 | -0.002 |
| FTE GPs per 1,000 patients | 0.099** | 0.051 |
| | -0.049 | -0.051 |
| FTE nurses per 1,000 patients | 0.044 | 0.052 |
| | -0.087 | -0.09 |
| FTE other direct staff per 1,000 patients | -0.218*** | -0.115 |
| | -0.069 | -0.072 |
| GP practice disease register - Atrial Fibrillation (in %) | 0.349*** | 0.336*** |
| | -0.046 | -0.048 |
| GP practice disease register - Asthma (in %) | 0.072*** | 0.030** |
| | -0.013 | -0.014 |
| GP practice disease register - Cancer (in %) | -0.016 | -0.058** |
| | -0.027 | -0.028 |
| GP practice disease register - Coronary heart disease (in %) | 0.039 | -0.117*** |
| | -0.033 | -0.036 |
| GP practice disease register - Chronic kidney disease (18+) (in %) | -0.025*** | -0.001 |
| | -0.009 | -0.009 |
| GP practice disease register - Chronic obstructive pulmonary disease (in %) | 0.064*** | 0.143*** |
| | -0.024 | -0.026 |
| GP practice disease register - Cardiovascular disease - primary prevention (30-74) (in %) | -0.015 | 0.017 |
| | -0.022 | -0.022 |
| GP practice disease register - Dementia (in %) | -0.118*** | -0.187*** |

| | | | |
|---|---|---|---|
| | -0.04 | -0.041 | |
| GP practice disease register - Depression (in %) | 0.002 | -0.009** | |
| | -0.004 | -0.004 | |
| GP practice disease register - Diabetes mellitus (17+) (in %) | 0.024** | 0.054*** | |
| | -0.01 | -0.011 | |
| GP practice disease register - Epilepsy (18+) (in %) | -0.128* | -0.388*** | |
| | -0.073 | -0.075 | |
| GP practice disease register - Heart failure (in %) | 0.054 | 0.015 | |
| | -0.043 | -0.046 | |
| GP practice disease register - Hypertension (in %) | 0.034*** | 0.014* | |
| | -0.008 | -0.008 | |
| GP practice disease register - Learning disability (in %) | 0.049 | 0.085 | |
| | -0.054 | -0.056 | |
| GP practice disease register - Mental health (in %) | -0.756*** | -0.488*** | |
| | -0.033 | -0.034 | |
| GP practice disease register - Obesity (in %) | 0.029*** | 0.014*** | |
| | -0.004 | -0.004 | |
| GP practice disease register - Osteoporosis (in %) | -0.055*** | -0.027 | |
| | -0.018 | -0.019 | |
| GP practice disease register - Peripheral arterial disease (in %) | -0.673*** | -0.199*** | |
| | -0.07 | -0.073 | |
| GP practice disease register - Palliative care (in %) | 0.061* | 0.051 | |
| | -0.034 | -0.036 | |
| GP practice disease register - Rheumatoid arthritis (16+) (in %) | -0.229*** | -0.062 | |
| | -0.067 | -0.07 | |
| GP practice disease register - Stroke and transient ischaemic attack (in %) | 0.192*** | 0.205*** | |
| | -0.051 | -0.053 | |
| Weighted Average Life expectancy at birth for GP practice | -0.074*** | 0.014 | |
| | -0.01 | -0.011 | |
| *CCG Fixed effects* | *No* | *Yes* | *No* |

| | | | |
|---|---|---|---|
| *Provider Fixed effects* | *Yes* | *Yes* | *Yes* |
| Constant | 15.719*** | 5.072*** | 11.369*** |
| | -1.639 | -1.714 | -1.354 |
| Observations | 13,888,084 | 13,888,084 | 14,452,415 |
| Number of Trusts | 127 | 127 | 127 |
| Within R2 | 0.144 | 0.144 | 0.144 |
| Between R2 | 0.175 | 0.141 | 0.175 |
| Overall R2 | 0.143 | 0.143 | 0.143 |

Standard errors in parentheses, *** p<0.01, ** p<0.05, * p<0.1
